# Creatine Kinase during Non-ST-Segment Elevation Acute Coronary Syndromes is Associated with Major Bleeding

**DOI:** 10.1101/2020.02.28.20029108

**Authors:** Lizzy M. Brewster, Jim D. Fernand

## Abstract

**Background:** It was recently reported that highly elevated plasma activity of the ADP-scavenging enzyme creatine kinase (CK), to >10 times the upper reference limit (URL), is independently associated with fatal or non-fatal bleeding during treatment for ST-segment elevation myocardial infarction (OR 2.6 [95% CI, 1.8 to 2.7]/log CK increase). Evidence indicates that CK attenuates ADP-dependent platelet aggregation. This study investigates whether moderately elevated CK in non-ST-segment elevation acute coronary syndromes (NSTE-ACS) is associated with major bleeding.

**Methods:** The Thrombolysis In Myocardial Ischemia (TIMI) 3B trial compared rt-PA (35 to 80 mg) with placebo, and early catheterization with conservative management in patients with NSTE-ACS. Main outcomes of the current study are the independent association of peak plasma CK (CKmax) with adjudicated fatal or non-fatal major bleeding (primary), and with combined major bleeding, stroke, and all-cause mortality (secondary) in multivariable binomial logistic regression analysis, with co-variables including age, sex, BMI, SBP, creatinine, and treatment assignment. Discrimination was assessed with C-statistics.

**Results:** The study included 1473 patients (66% men, 80% white, mean age 59 y, SE 0.3). CKmax ranged between 15 and 19045 IU/L (mean (SE), 450(24) IU/L; i.e. 2 times URL). Major bleeding occurred in 2.0% (mean age 65(1.3) y; mean CKmax 1015(318) IU/L; 6 times URL), and the combined outcome in 4.3% of the patients, adjusted OR per log CK increase respectively 3.1 [1.6 to 5.8] for major bleeding, and 3.9 [2.5 to 6.1] for the combined outcome; C-index 0.8 for both outcomes.

**Discussion:** The presented data add to the existing evidence that proportionate to its plasma activity, the ADP-binding enzyme CK is strongly and independently associated with non-fatal and fatal major bleeding during ACS treatment. CK might increase the accuracy of prediction models for major bleeding in patients treated with antithrombotic or thrombolytic drugs for ACS.

**ClinicalTrials.gov identifier:** NCT00000472

## Background

Major bleeding is the most common non-cardiac cause of mortality during treatment of non-ST-segment elevation acute coronary syndromes (NSTE-ACS). Therefore, it is important to identify patients at risk for hemorrhagic complications during the management of ACS.^1-4^ Evidence indicates that the ADP-binding enzyme creatine kinase (CK) inhibits ADP-dependent platelet activation and subsequent aggregation (**Figure 1**).^5-8^ During myocardial ischemia and infarction, CK and other cell constituents such as myoglobin and troponin enter plasma along with smaller molecules including phosphocreatine.^6-8^ Extracellular CK is thought to reduce ADP and ADP-dependent platelet activation through its scavenging action on ADP, or through conversion into ATP, catalyzing the reaction:^6-8^

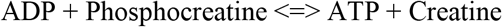

**Figure 1.**
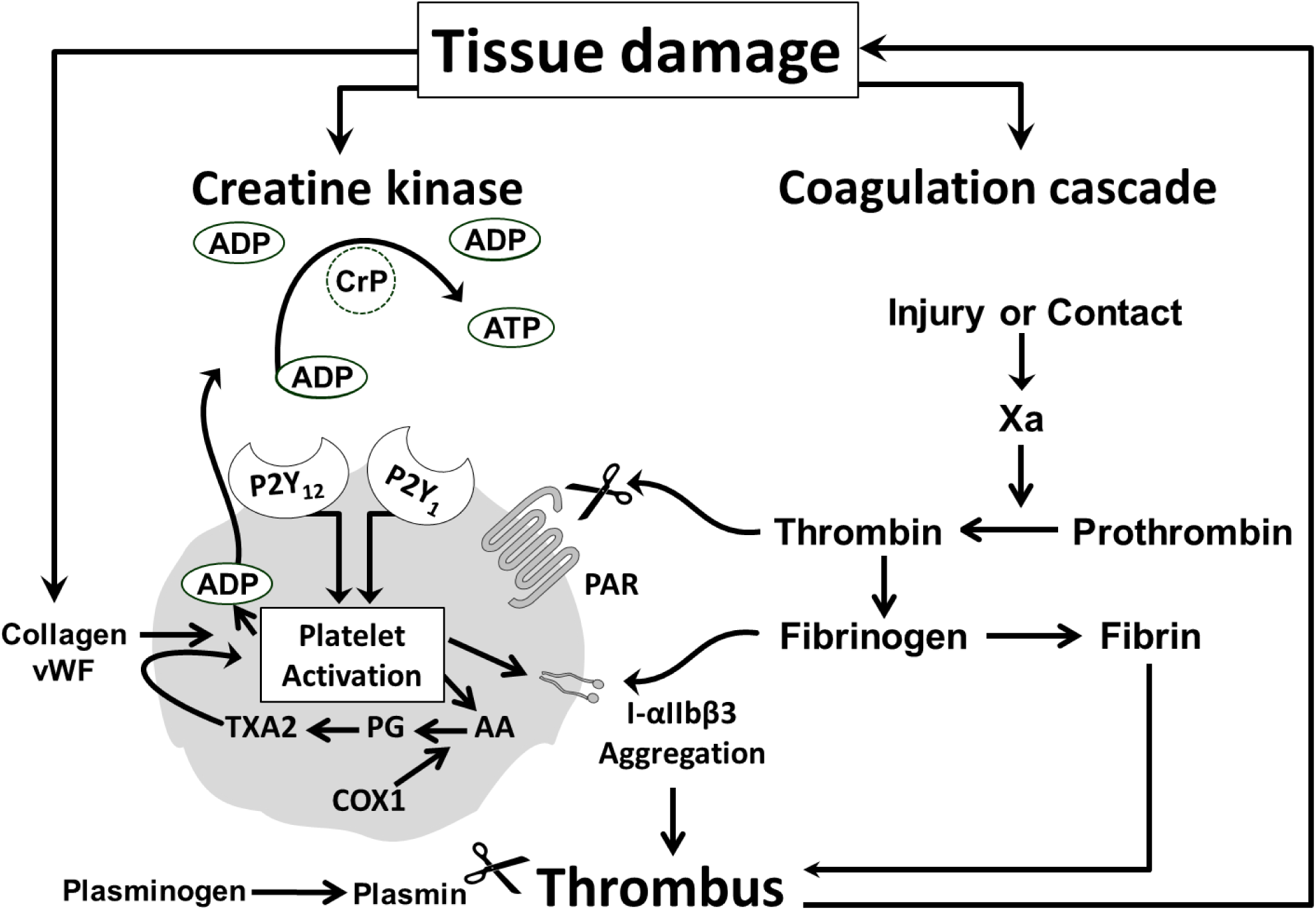
The ADP-binding enzyme creatine kinase and inhibition of platelet activation. Central to the development of acute coronary syndromes (ACS) is plaque rupture or erosion resulting in platelet activation, aggregation, and thrombosis.^1-5^ Creatine kinase (CK) was recently proposed to be an essential part of the counterregulatory mechanism of endothelial thromboresistance that limit platelet activation.^6,7^ After plaque rupture platelets adhere to the injured vessel wall and undergo activation through collagen and von Willebrand factor (vWF). Activated platelets release substances including ADP and thromboxane A2 (TXA2) synthesized from arachidonic acid (AA) through prostaglandin (PG) catalyzed by (COX1). ADP and TXA2 amplify the response to injury and produce sustained platelet aggregation. Furthermore, thrombin generated by tissue damage activates platelets’ protease activated receptors (PAR), leading to the generation of more thrombin on the platelets’ surface, further platelet activation, conversion of fibrinogen to fibrin, and further stabilization of the platelet-fibrin clot. Fibrin is degraded by plasmin, formed by plasminogen and (recombinant) tissue-type plasminogen activator. ADP is considered to be central to platelet activation. ADP-stimulation of the P2Y1 receptor activates phospholipase C resulting in weak, transient platelet aggregation. Activation of P2Y12 receptor results in the activation of glycoprotein receptors IIb/IIIa (integrin (I)-αIIbβ3) and firm platelet aggregation.^5,6^ CK strongly binds ADP and reduces platelet aggregation in the presence of phosphocreatine (CrP).^7,8^ The enzyme might act in synergy with antithrombotic and fibrinolytic drugs that act on Factor Xa, Thrombin, PAR, COX1, P2Y12, I-αIIbβ3, plasminogen or other factors^1,2^ to increase bleeding risk in ACS.^6^ (Modified after Brewster and Fernand, 2019).^6^

In line with this, it was recently reported that peak plasma CK (CKmax) during admission is strongly and independently associated with bleeding after ST-elevation myocardial infarction (STEMI), with an adjusted odds ratio (OR) per log CK increase of 2.6 [95% CI, 1.8 to 3.7] for fatal/non-fatal bleeding, and 3.1 [2.2 to 4.4] for bleeding/all-cause mortality.^6^ As the effect of CK on ADP-dependent platelet aggregation is thought to act in synergy with antithrombotic and thrombolytic therapy,^6^ lower levels of CK might also be associated with bleeding. Therefore, it is studied whether moderately elevated CK during NSTE-ACS is independently predictive of major bleeding.

## Methods

### Ethical approval

The study protocol was approved by the Review Board of the Research Ethics Committee of the Radboud University Nijmegen Medical Centre on April 30, 2019 (registration number 2019-5356), and by the US National Heart, Lung, and Blood Institute (RMDA V02 1d20120806) on May 6, 2019.

### The TIMI 3 trial

Data were used of the Thrombolysis In Myocardial Ischemia Trial 3 (TIMI 3) where CK was routinely estimated during admission in all patients.^9,10^ As reported previously,^9,10^ the TIMI 3 study consisted of two separate multicenter, randomized controlled double blind clinical trials designed to assess the effects of recombinant tissue plasminogen activator (rt-PA) vs. placebo added to conventional therapy (including intravenous heparin and oral aspirin) on the coronary culprit lesion (TIMI 3A, N=391), and on clinical outcomes (TIMI 3B, N=1473) in comparably selected and treated patients with unstable angina or non-ST-segment elevation myocardial infarction. TIMI 3B further compared management with early cardiac catheterization (and percutaneous transluminal coronary angioplasty or coronary artery bypass surgery, if appropriate) at 18 to 48 hours (h) after presentation (Invasive Strategy), versus angiography with revascularization only if the patient had spontaneous or provokable ischemia (Conservative Strategy). The TIMI 3A study recruited participants aged 21 to 76 y, and TIMI 3B aged 21 to 79 y, in respectively 15 and 25 participating centers across the United States of America and Canada. All patients had given written informed consent to participate in the study.^9,10^ Patients were required to have chest discomfort at rest suggestive of myocardial ischemia, lasting greater than 5 minutes but less than 6 hours, which occurred within 12 hours (24 hours for TIMI 3B) of the time of enrollment. Not eligible for inclusion were patients who underwent bypass surgery, had experienced MI within the preceding 21 days (excluding within 12 hours before enrollment), had new, persistent ST-segment elevation, had PTCA in the past 6 months, had contraindication for thrombolytic therapy, used oral anticoagulant therapy, had any history of cerebrovascular disease or transient ischemic attack, or advanced illness including malignancies, hepatic, renal disorders, among other criteria. Standard clinical care included heparin given as a bolus of 5000 IU iv followed by an infusion of 1000 IU/h to reach aPTT 1.5 to 2.0 times normal, and anti-ischemic therapy with a beta-adrenergic blocker, a calcium channel blocker, and a nitrate preparation. Aspirin was not given routinely in TIMI 3A and 325 mg daily from Day 2 in TIMI 3B. The primary comparison was rt-PA vs. placebo (blinded), with a primary end point of improvement in perfusion or stenosis in TIMI 3A. In TIMI 3B, the secondary comparison was Invasive vs. Conservative Strategy (unblinded), with a combined primary endpoint of reduction in death, (subsequent) myocardial infarction, or either spontaneous or provokable ischemia within or at 6 weeks after study drug infusion. The main secondary evaluation was at Day 21 after submission.^9,10^

### Prevention and Management of Hemorrhagic Complications

Taken the high bleeding rate in the TIMI 2 trial into account (investigator-reported bleeding in 58.4% of the patients),^6,11^ elaborate measures were taken in TIMI 3 to prevent and treat hemorrhage, such as more stringent contraindications for thrombolytic therapy compared to TIMI 2.^6,9-11^ While in TIMI 2, 100 to 150 mg rt-PA was used, rt-PA dose was reduced in TIMI 3 to 0.8 mg/kg, with a maximum of 80 mg. aPTT-guided heparin dose adjustments were part of the study protocol, and relating to the high number of puncture bleeds in TIMI 2,^6,11^ heparin doses were routinely reduced 4 hours prior to the first arterial puncture for the arteriogram. Major vascular catheter entry sites were minimized and protected with sheath-introducers to tamponade the site for approximately 24 hours. Vascular puncture sites were treated with compression dressings, and sites of vascular entry were checked every two hours during the acute study period. Other measures included complete blood count at least every 12 hours during the first 24 hours after the intervention, and testing of stool, urine and vomitus specimens for occult blood. Frequent clinical assessments and examinations were advised to assess potential internal hemorrhage. If bleeding occurred, management was highly protocolized, and specific therapy was given according to site of bleeding, such as antacids and histamine 2 receptor blockers for upper gastrointestinal bleeding. Finally, life-threatening hemorrhagic complications or any were reported immediately to the Coordinating Center and the Program Office.^9,10^

### Classification of Hemorrhagic Events

Bleeding was classified by the Hemorrhagic Event Review Committee as described previously,^11^ as “major” when a decrease in hemoglobin of >50 g/L (>3.1 mmol/L), intracranial bleeding, or cardiac tamponade was present, or with death from hemorrhage; as “minor” if a hemoglobin reduction 30 to 50 g/L (1.9 to 3.1 mmol/L) from an identified bleeding site was present, or if the patient had spontaneous macroscopic hematuria, hemoptysis, or hematemesis; and as “loss-no-site” if the bleeding site was unknown, but hemoglobin reduction was 40 to 50 g/L (2.5 to 3.1 mmol/L). Transfusion of one unit of packed cells or whole blood was counted as 10 g/L or hematocrit reduction of 3%.^9-11^

### Laboratory studies

Total CK activity was estimated at the study centers, collected at baseline (pre-treatment), during the first 3 days, at 4, 12, 24, 48, and 72 hours, and thereafter when indicated. The upper reference limit (URL) as defined by the local laboratory was recorded. CK twice the URL was considered evidence of infarction. Fibrinogen levels and fibrin(ogen) degradation products, were collected at baseline (pre-treatment), and at 50 min, and at 12, 24, 48, 96 hours after rt-PA infusion, and determined in a central Coagulation Core Laboratory, using standardized methods as described previously.^9,10^ Platelet count was determined locally at the participating centers.

### Main outcomes

The main outcomes of this analysis are the independent association of peak CK activity with adjudicated major bleeding (primary); and with combined adjudicated major bleeding, stroke, and all-cause mortality (ACM) during hospitalization (secondary).

### Sample size considerations

The probability P of events at the mean value of the CK (and other variables) was conservatively estimated to be 0.10. Based on the previous report,^6^ sample size was estimated for two levels of the OR of disease, corresponding to an increase of one standard deviation from the mean value of CK, given the mean values of the remaining variables; at OR 1.2, with at least 634 participants needed; or at OR 2.0, with at least 226 needed for this analysis with a one-tailed alpha of 5% and 1 – beta of 90. A second consideration was to include a minimum of 10 events of the least frequent outcome per parameter to fit the model to avoid over or under estimated variances.^12^

### Data analyses

Data of the TIMI 3B study are analyzed. Demographic characteristics are reported by bleeding status. Parametric versus nonparametric statistical methods are used where appropriate. CK data are known to be highly skewed to the right and log transformation to the base of 10 is used in regression analysis.^6^ Mean as well as peak CK during admission are assessed (primary variable), as crude values, and standardized by the local upper reference limit (URL). In addition, time to CKmax, and time to, severity, and location of the first adjudicated bleeding are noted. Major bleeding is associated with CKmax presented in categories, and as a continuous measure in univariable and multivariable logistic regression analysis, to assess whether log CKmax is independently predictive of fatal or non-fatal major bleeding (primary), or combined major bleeding, stroke, and all-cause mortality (ACM, secondary), at Day 21 after hospital admission. Five covariables are chosen a priori, sex, age, body mass index (BMI, body weight in kg divided by the squared height in meters), history of diabetes, and systolic blood pressure (SBP) at admission. Other variables are considered based on statistical and clinical criteria and bleeding risk scores^1-6;9-11^ to find the most parsimonious model that explains the data, using forced entry and bootstrapping for internal validation. Global model fit is based on Bayes information criterion (BIC), and calibration included Hosmer-Lemeshow statistics. Main subgroup analysis is by rt-PA treatment assignment vs. placebo. Sensitivity analysis include modelling with different relevant covariables and by peak CK at 24h. Model discrimination is assessed with C-statistics.^13^ Missing data are not imputed, and p-values are not reported for study outcomes.^14^ Where relevant, statistical uncertainty surrounding the estimates is communicated through 95% confidence intervals (CI) given between square brackets, with uncertainty regarding the external validity discussed. Data in parentheses are SE unless indicated otherwise. Statistical analyses are performed with the SPSS statistical software package for Windows version 25.0 (SPSS Inc, Chicago, Ill, USA).

## Results

### Baseline Characteristics and Assigned Treatment

TIMI 3B patients were predominantly men (66.3%) and white (79.7%), with a mean age of 58.8 (0.3) years (**Table 1**). Patients were randomized to rt-PA (49.4%) or placebo (50.6%), and to the invasive strategy (49.8%) vs. the conservative strategy (50.2%). Median duration of hospitalization was 11 days (interquartile range, IQR 7 to 14 days). During the first 5 days of hospitalization, nearly all patients were given heparin (99.7%) and aspirin (99.5%), with a median total daily dose (IQR) of 17 519 U USP (IQR 13 600 to 21 320) for heparin, and 260 mg (195 to 284) for aspirin.

**Table 1.** Patients’ Characteristics. Patients by adjudicated (adj.), major bleeding (primary outcome) vs. other or no bleeding. Data are counts, mean (SD); or median (IQR), rounded to whole numbers where applicable.*At admission (baseline), †Derived from days. BMI, body mass index, (F)H/O, (family) history of; MI, myocardial infarction; CAD, coronary artery disease; SBP, systolic blood pressure, HB, hemoglobin, HT, Hematocrit, WBC, white blood cell; rt-PA, recombinant tissue-type plasminogen activator; FDP, fibrin(ogen) degradation products; d, day; CKmax, peak plasma creatine kinase during admission; upper reference limit (URL); h, hour; n.a, not applicable. Anticoagulant drug use was an exclusion criterion.

| Clinical Parameters | Bleeding |  |  |
| --- | --- | --- | --- |
|  | None | Non-Major | Adj. Major |
| N | 1345 | 98 | 30 |
| Men (%) | 67 | 57 | 70 |
| Age (y) | 58 (0.3) | 61 (0.1) | 65 (1.3) |
| Ancestry (white,%) | 79 | 90 | 80 |
| BMI (kg/m<sup>2</sup>) | 28 (0.1) | 27 (0.5) | 27 (0.8) |
| H/O Cigarette smoking | 69 | 68 | 69 |
| H/O Hypertension (%) | 42 | 43 | 36 |
| H/O Diabetes mellitus (%) | 15 | 11 | 19 |
| H/O Previous angina | 85 | 92 | 87 |
| History of MI (%) | 41 | 34 | 21 |
| FH/O CAD | 39 | 35 | 23 |
| On diuretics at adm. | 16 | 20 | 30 |
| Aspirin within 24 h before adm. | 48 | 43 | 45 |
| Aspirin dose before adm. | 325 (0 to 325) | 325 (0 to 325) | 325 (0 to 325) |
| R/ Hypercholesterolemia | 20 | 15 | 7 |
| SBP (mm Hg)* | 132 (0.6) | 132 (1.8) | 129 (2.8) |
| Platelet count (·10<sup>9</sup>/L)* | 264 (2.0) | 275 (9.2) | 269 (14.2) |
| HB (mmol/L)* | 8.8 (0.0) | 8.6 (0.1) | 9.3 (0.7) |
| HT (%)* | 41.5 (0.1) | 40.9 (0.5) | 40.5 (1.4) |
| aPTT >1.5 URL (%) at adm. | 12 | 11 | 13 |
| WBC count (·10<sup>9</sup>/L)* | 8.7 | 9.0 | 10.6 |
| Creatinine (micromol/L)* | 97 (80 to 115) | 88 (80 to 106) | 97 (71 to 117) |
| Invasive treatment arm (%) | 49 | 61 | 62 |
| rt-PA (%) | 48 | 57 | 73 |
| Fibrinogen 12h (g/L) | 2.7 (0.0) | 2.6 (0.1) | 2.4 (0.1) |
| FDP 12h min (mg/L) | 0.22 (0.11) | 0.24 (0.24) | 1.20 (0.8) |
| Heparin dose d. 1 to 5 (U/d) | 17 620 (169) | 16 835 (565) | 15 263 (1 402) |
| Aspirin dose d. 1 to 5 (mg/d) | 240 (2.4) | 234 (10.7) | 181 (24.4) |
| HB d. 1 to 5 (mmol/L) | 12.9 (0.0) | 12.0 (0.2) | 11.6 (0.3) |
| Platelet count d. 1 to 5 ($\cdot 10^9/L$) | 238 (1.9) | 242 (7.6) | 202 (12.3) |
| CKmax (IU/L) | 149 (82 to 424) | 180 (83 to 436) | 390 (174 to 1104) |
| CKmax (times URL) | 0.7 (0.4 to 2.0) | 0.9 (0.4 to 2.0) | 1.6 (0.9 to 5.7) |
| CKmax <200 IU/L/<URL (%) | 60/59 | 55/53 | 30/40 |
| Time to peak CK (h) | 4 (0 to 24) | 4 (12 to 48) | 4 (24 to 72) |
| Time to bleeding (h) | n.a. | 96 (72 to 120) | 72 (48 to 144) |

### Trial Outcomes

TIMI3 trial endpoints by treatment arm have been reported previously.^9,10^ In summary, in TIMI 3A, of all culprit lesions, 25% with rt-PA vs. 19% with placebo achieved the primary endpoint of measurable improvement. A thrombus was present in the culprit lesion in 107 patients (35%), and in these patients, substantial improvement occurred in 36% of the patients with rt-PA vs. 15% with placebo. However, in the TIMI 3B study, the incidence of death or non-fatal infarction did not differ after one year by strategy assignment, although fewer patients in the early invasive strategy group underwent repeat hospital admission.^9,10^

### CK

CK was estimated at 18 different time points, in total 9 215 times in 1 470 patients during the hospital stay, from the day of admission to day 17. Plasma CK, missing in 4 patients, ranged between <10 to 19 045 IU/L. Mean CK per patient was 221(9) IU/L, with a mean URL of 212 IU/L. CKmax ranged between 15 and 19 045 IU/L (mean 450(24); median 152, IQR 82 to 433), a mean of 2.2(0.1) times the URL (**Figure 2, Panel A and B**). Most patients (58%) had a peak CK level below the URL, and only 25% of the patients had a CK greater than twice normal; with the highest CK activity on the first day in most patients (**Figure 2. Panel C**).

**Figure 2.**
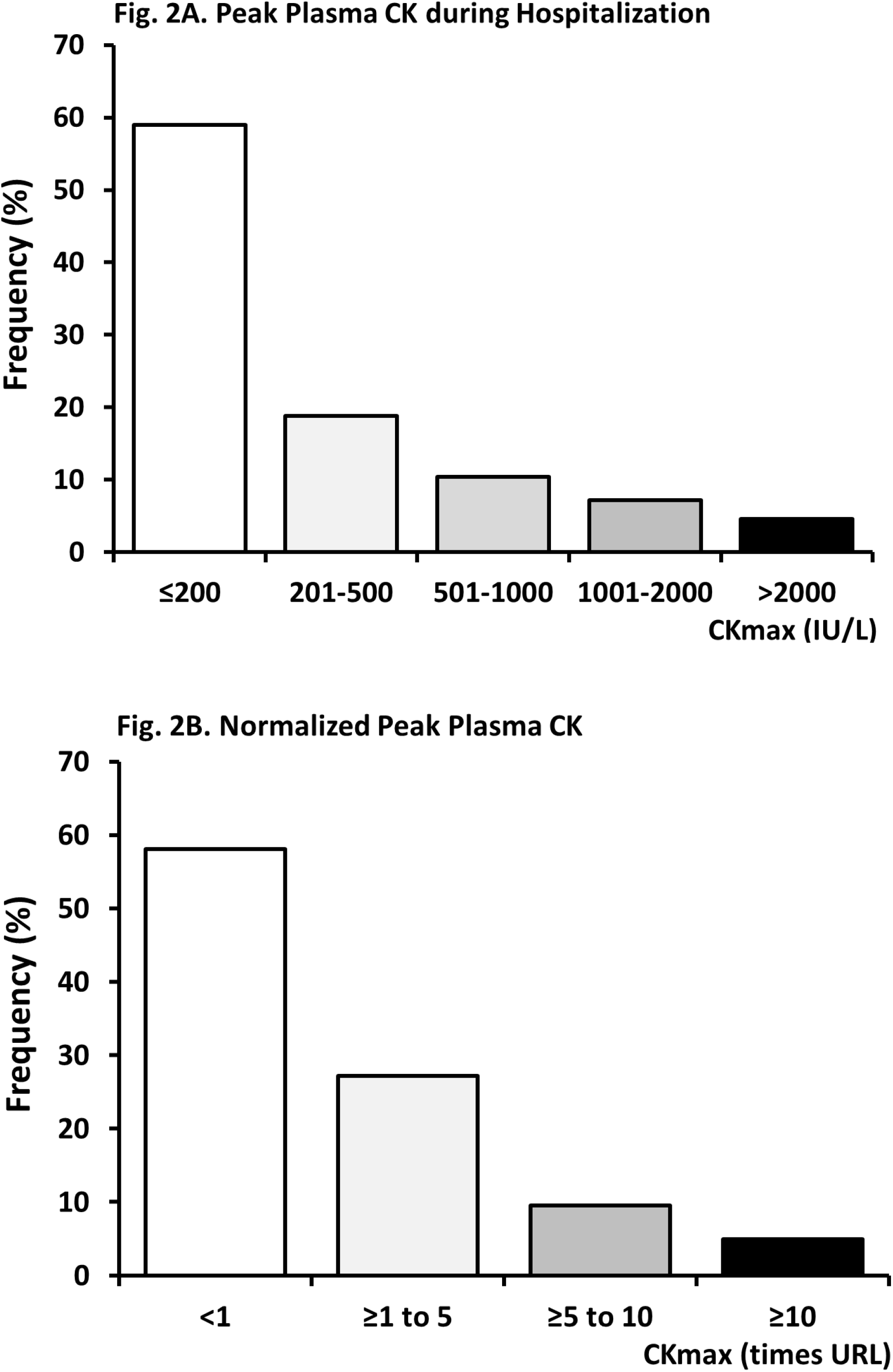
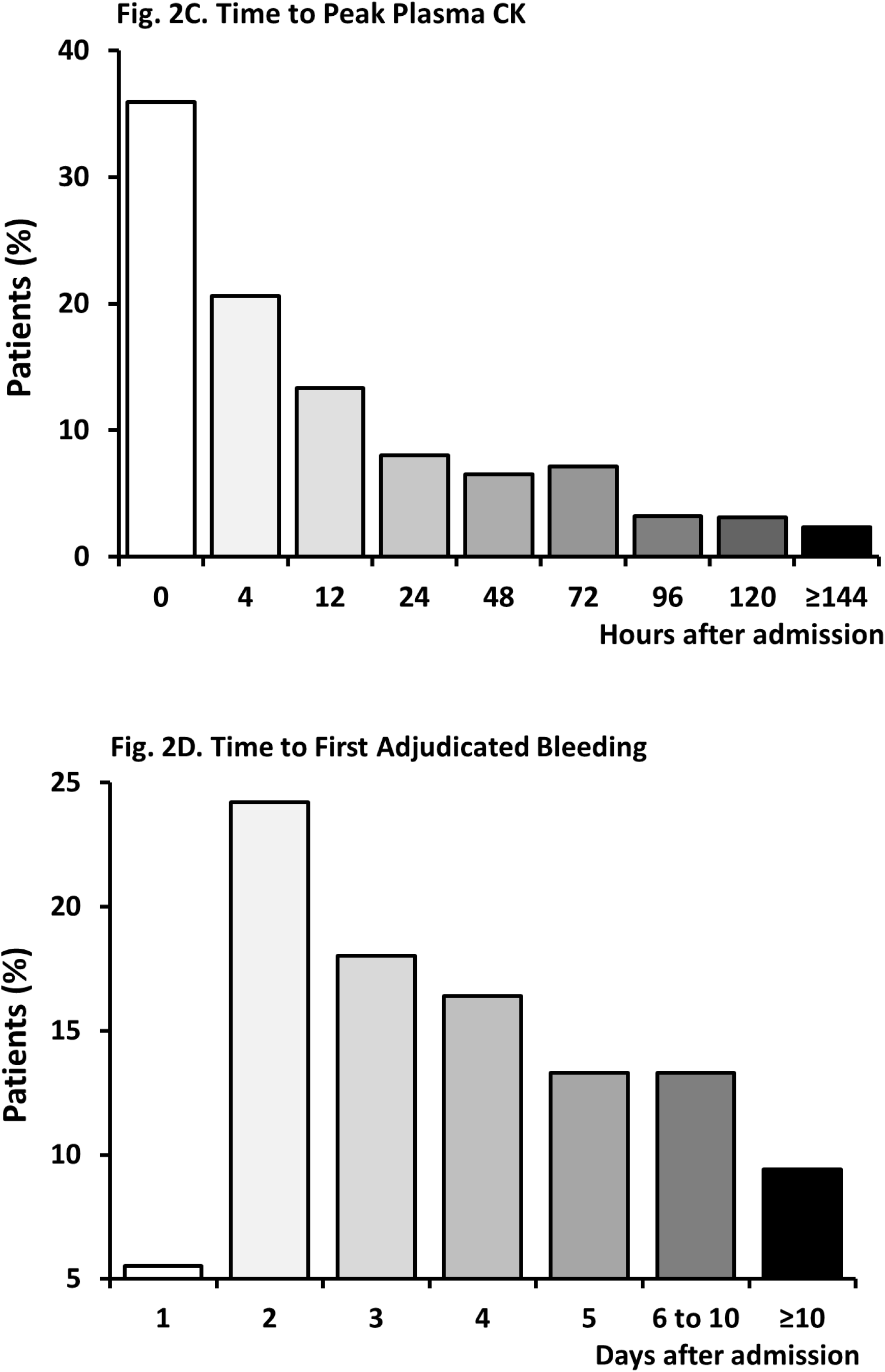
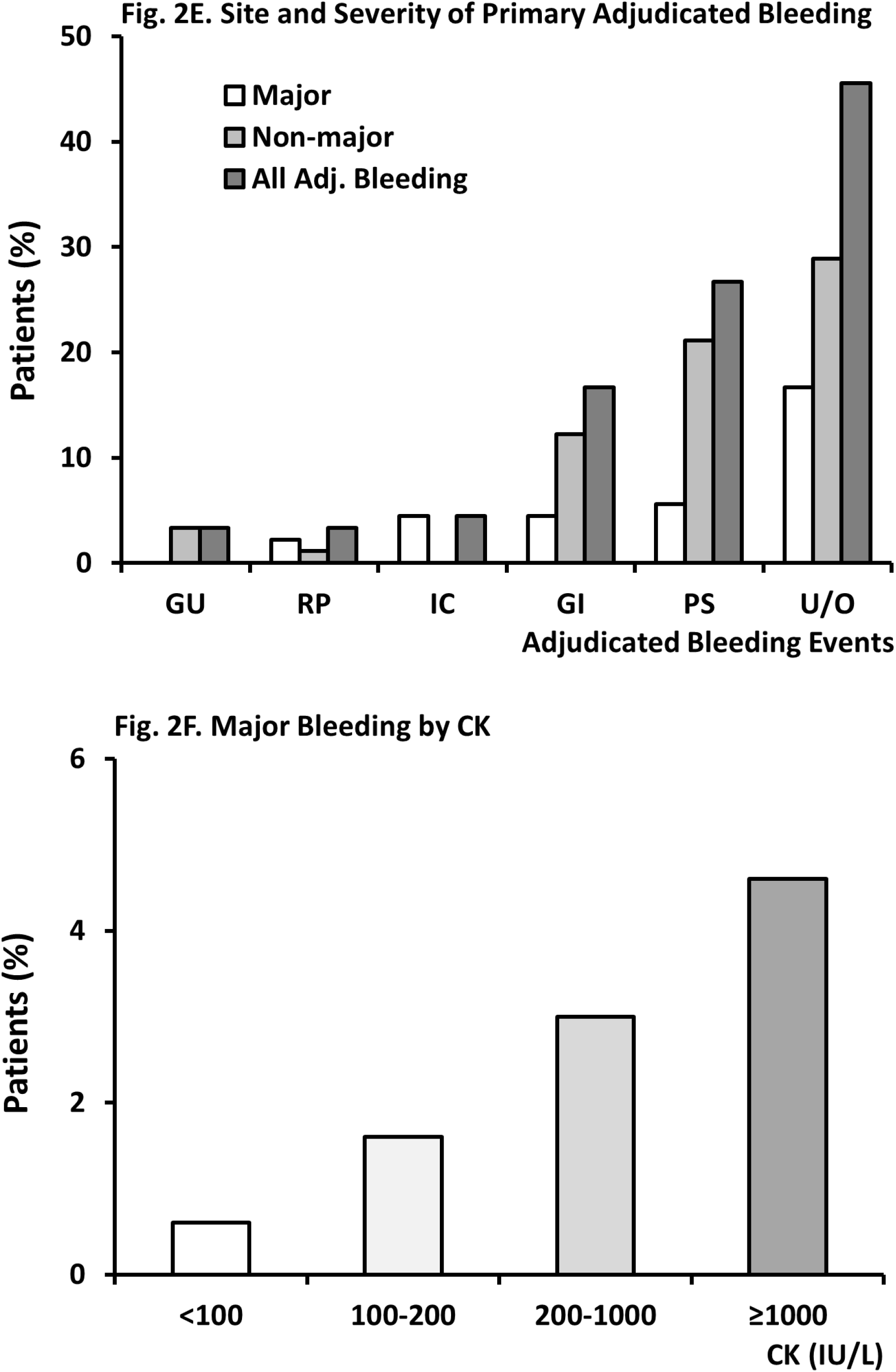
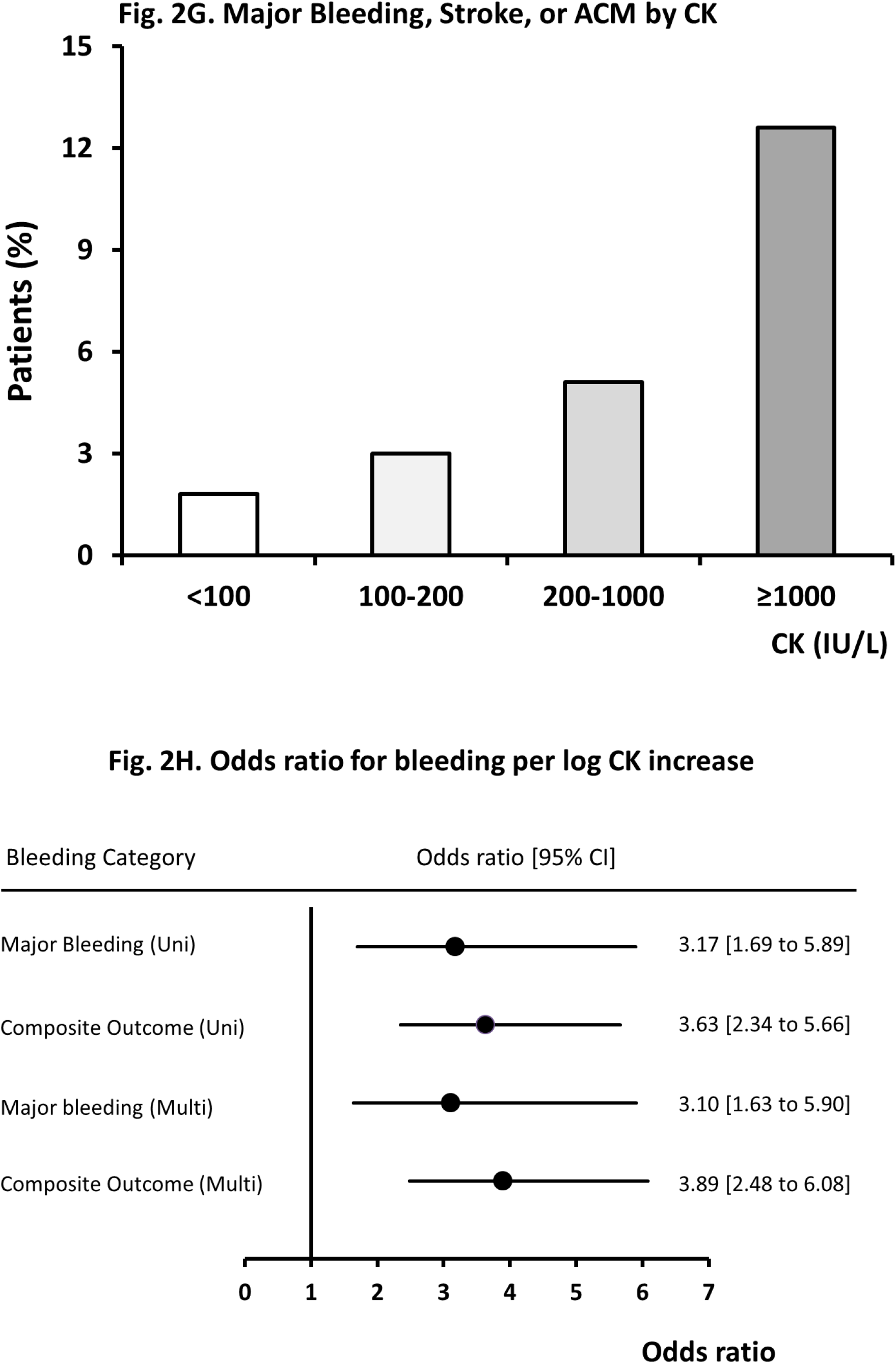
The Association between CK and Bleeding. The association between creatine kinase (CK) and bleeding. Panel A (IU/L) and B (CK normalized for the upper reference limit, URL) show CK activity below URL in the majority of patients, as expected with non-ST-segment elevation acute coronary syndromes. Panel C and D depict the time to peak CK value (day 1) vs. time to the first adjudicated bleeding (peak at Day2). Panel E shows the site and severity of adjudicated bleeding (with surgical bleeding classified as “other”), and Panel G and H the primary and secondary outcome by CK level, with crude CK levels and in univariable and multivariable logistic regression analysis by CK adjusted for URL (panel H). GU, RP, IC, GI, PS, U/O are respectively, genitourinary, retroperitoneal, intracranial, gastrointestinal, puncture site (vascular access site-related), or unknown/other. Panel A-B. Peak plasma CK activity Panel C-D. Timing of peak CK and bleeding Panel E-F. Bleeding events by Site and CK Panel G-H. Composite outcomes by CK (Univariable and Multivariable)

### Hemorrhagic complications

Bleeding, reported in 128 patients (8.7%), was most frequent on the second day after admission (24.2%, **Figure 2, Panel D**). Adjudicated bleeding events were registered in 90 patients, 30 (33.3%) with major bleeding, 43 (41.1%) minor bleeding, and 23 (25.6%) loss-no-site. Primary bleeding locations are depicted by severity in **Figure 2, Panel E**, with surgery-related bleeding (11 patients) classified as “other”. Stroke (hemorrhagic and non-hemorrhagic) was reported in 18 patients. Thirty patients died during hospitalization (2.1%), of whom 9 (30%) had a major bleeding or stroke. Seven out of 30 patients (23.3%) with a major bleeding died, and 64 patients (4.3%) had a major bleeding, stroke or ACM.

Mean CKmax in patients without major bleeding was 439(23) IU/L, (2.2*URL, SE 0.1), vs 1 015(319) IU/L, (5.6*URL, SE 2.4) with major bleeding. The association between CK and major bleeding included “normal” CK levels (**Figure 2, Panel F and G**). In univariable binary logistic regression analysis, the OR of fatal or non-fatal major bleeding was 3.2 [1.7 to 6.0]/log CKmax increase. For the composite outcome major bleeding, stroke, or hospital death, mean CKmax was 1 177(231), (6.1*URL, SE 1.5); mean age 64(1.0) years, OR 3.6 [2.3 to 5.7]. Sex, age, ancestry (white vs. non-white), BMI, a history of diabetes or hypertension, baseline SBP, baseline laboratory values (e.g. hematocrit, creatinine, platelet count), and treatment assignment (conservative vs. invasive and rt-PA vs. placebo), were further considered as predictors of the main primary and secondary outcomes. Data on congestive heart failure and heart rate at admission were not available. Multivariable binary logistic regression analysis suggested an independent association between peak CK and major bleeding as well as the composite outcome, with per log CK increase a 3-fold increase in odds for non-fatal or fatal major bleeding and a nearly 4-fold increase in combined major bleeding, stroke, or ACM, compared to the absence of these outcomes (**Table 2 A and B; and Figure 2 Panel H**). The C-index of the final models was 0.8 (**Figure 3**). Although the number of events in the subgroups were too small to build a stable statistical model, the data indicated that the association between CK and major bleeding was similar in size and direction in patients who received placebo (respectively 6.12 [1.70 to 21.99], n=703); vs. rt-PA 2.27 [1.08 to 4.77], n=667).

**Table 2A.** Modelling of the Association between CK and Major Bleeding. Modelling of the association between creatine kinase (CK) and major bleeding. *Women vs. men; **†**non-White vs. White; BMI, body mass index; SBP, systolic blood pressure; rt-PA, recombinant tissue-type plasminogen activator. Parameter units are as in Table 1. Baseline platelet count, and Fibrinogen and Fibrin (ogen) degradation products at 50 min or 12 h after start therapy were not associated with major bleeding (OR 1; data not shown). Multivar., multivariable full and restricted models. Model parameters full (restricted) model: omnibus test of model coefficients Chi-square 34.2 (32.4) df 6 (3); –2 Log likelihood 251 (253); BIC 270 (262) Cox and Snell R square. 0.02 (0.02); Nagelkerke R square 0.13 (0.12); Hosmer Lemeshow Chi square 6.2 (9.8), df 8 (8); classification overall percentage 98.0 (98.0), n=1465 (1465) complete, unimputed cases. Bootstrapping OR was identical to the depicted outcomes (data not shown). **‡**OR Age per 10 years (under the assumption of linearity) is 2.27 [1.42 to 3.63]. Including diuretic use at admission did not affect the final model. Substituting (log) CKmax during admission with (log) peak CK on day 1 yielded OR 2.64 [1.56 to 4.47] in the final model.

| Univariable | OR |  | Confidence Interval |
| --- | --- | --- | --- |
| **CK** | **3.17** |  | **[1.69 to 5.89]** |
| **Sex\*** | 0.84 |  | [0.38 to 1.85] |
| **Age** | **1.08** |  | **[1.03 to 1.13]** |
| **BMI** | 0.95 |  | [0.87 to 1.02] |
| **Ancestry†** | 0.98 |  | [0.40 to 2.42] |
| **Diabetes Mellitus** | 1.33 |  | [0.40 to 4.46] |
| **Hypertension** | 0.70 |  | [0.33 to 1.50] |
| **Baseline SBP** | 1.00 |  | [0.98 to 1.01] |
| **Baseline Hematocrit** | 0.95 |  | [0.88 to 1.04] |
| **Baseline Creatinine** | 0.93 |  | [0.81 to 1.07] |
| **rt-PA vs. Placebo** | **3.29** |  | **[1.40 to 7.74]** |
| **Invasive vs. Conservative** | 1.64 |  | [0.77 to 3.49] |

| Multivar. (Full ) | B | SE | Wald | OR | Confidence Interval |  |
| --- | --- | --- | --- | --- | --- | --- |
| **CK** | 1.10 | 0.33 | 11.14 | **3.01** | **1.58** | **5.76** |
| **Sex** | − 0.27 | 0.42 | 0.40 | 0.77 | 0.34 | 1.75 |
| **Age** | 0.08 | 0.03 | 11.50 | **1.09** | **1.04** | **1.14** |
| **Diabetes Mellitus** | 0.34 | 0.64 | 0.29 | 1.41 | 0.40 | 4.93 |
| **rt-PA vs. Placebo** | 1.23 | 0.44 | 7.80 | **3.43** | **1.44** | **8.15** |
| **Invasive vs. Conservative** | 0.44 | 0.39 | 1.24 | 1.55 | 0.72 | 3.34 |
| **Constant** | − 10.19 | 1.66 | 37.50 | 0.00 | - | - |

| Multivar. (Restricted) |  |  |  |  |  |  |
| --- | --- | --- | --- | --- | --- | --- |
| **CK** | 1.13 | 0.33 | 11.92 | **3.10** | **1.63** | **5.90** |
| **Age‡** | 0.08 | 0.02 | 11.24 | **1.09** | **1.04** | **1.14** |
| **rt-PA vs. Placebo** | 1.24 | 0.44 | 7.88 | **3.45** | **1.45** | **8.18** |
| **Constant** | − 9.90 | 1.64 | 36.33 | 0.00 | - | - |

**Table 2B.**
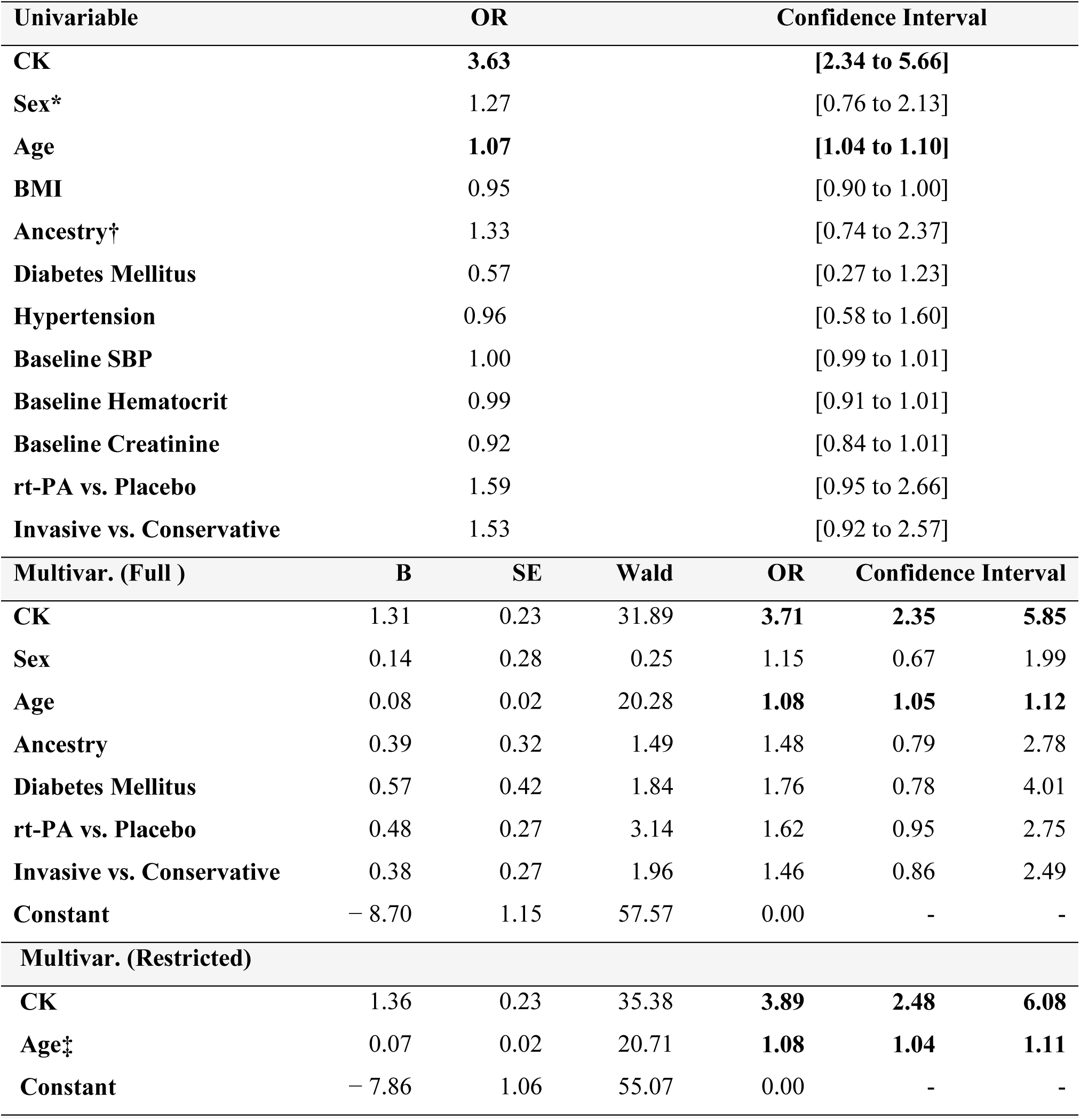
Modelling of the Association between CK and Major Bleeding, Stroke, or Death. Modelling of the association between (log) creatine kinase (CK) and the composite outcome (major bleeding, stroke, and death). *Women vs. men; **†**non-White vs. White; BMI, body mass index; SBP, systolic blood pressure; rt-PA, recombinant tissue-type plasminogen activator. Parameter units are as in Table 1. Baseline platelet count, and Fibrinogen and Fibrin(ogen) degradation products at 50 min or 12 h after start therapy were not associated with the composite outcome (OR 1; data not shown). Multivar., multivariable full and restricted models. Model parameters full (restricted) model: omnibus test of model coefficients Chi-square 63.1 (56.3) df 7 (2); –2 Log likelihood 457 (470); BIC 479 (476); Cox and Snell R square. 0.04 (0.04); Nagelkerke R square 0.14 (0.13); Hosmer Lemeshow Chi square 6.6 (10.2), df 8 (8); classification overall percentage 95.7 (95.6), n=1465 (1469) complete, unimputed cases. Bootstrapping OR was identical to the depicted outcomes (data not shown). ^**‡**^OR Age per 10y is 2.10 [1.53 to 2.87]). Including diuretic use at admission did not affect the final model. Substituting (log) CKmax during admission with (log) peak CK on day 1 yielded OR 2.99 [2.07 to 4.30] for CK in the final model.

**Figure 3.**
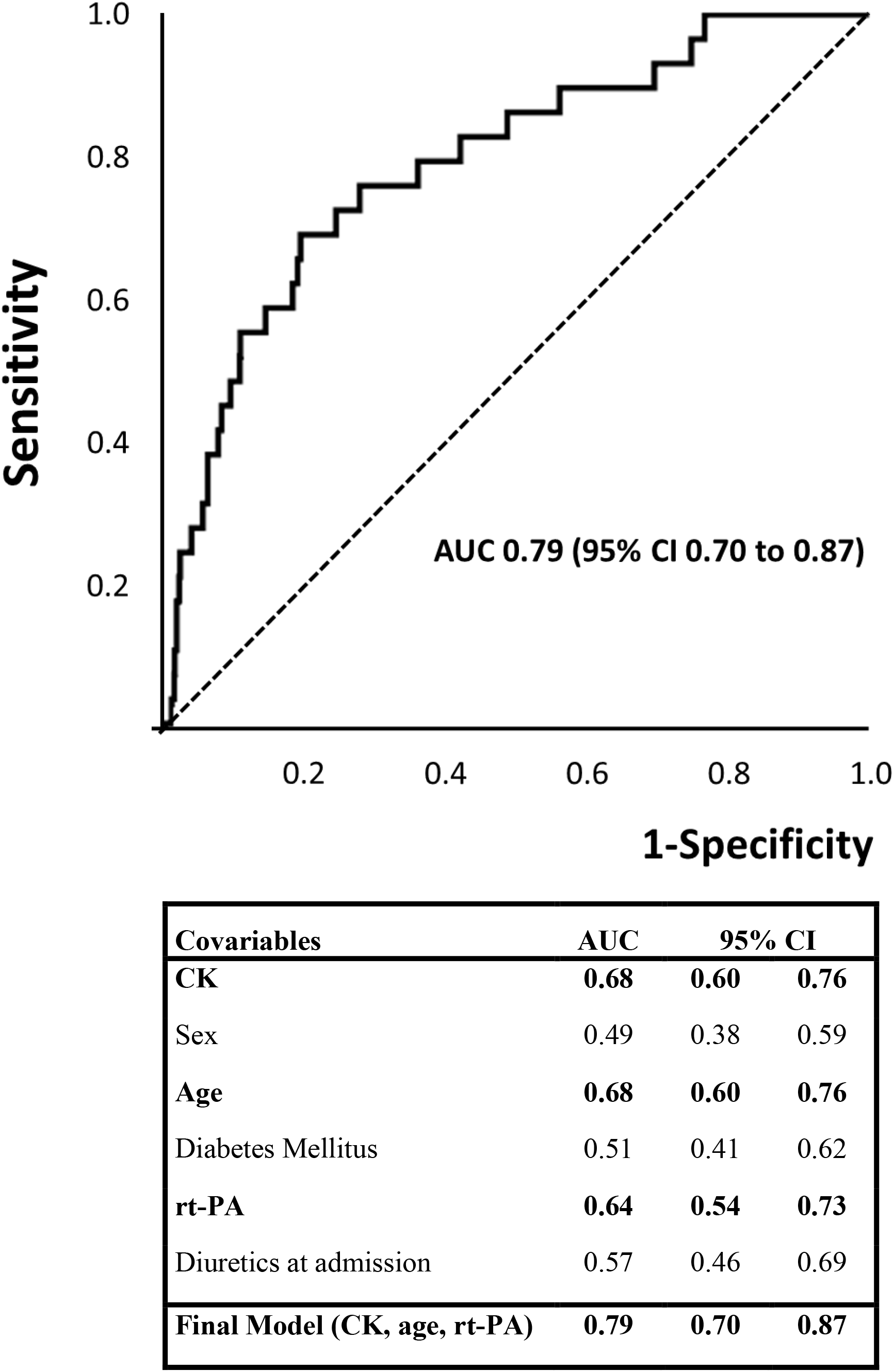
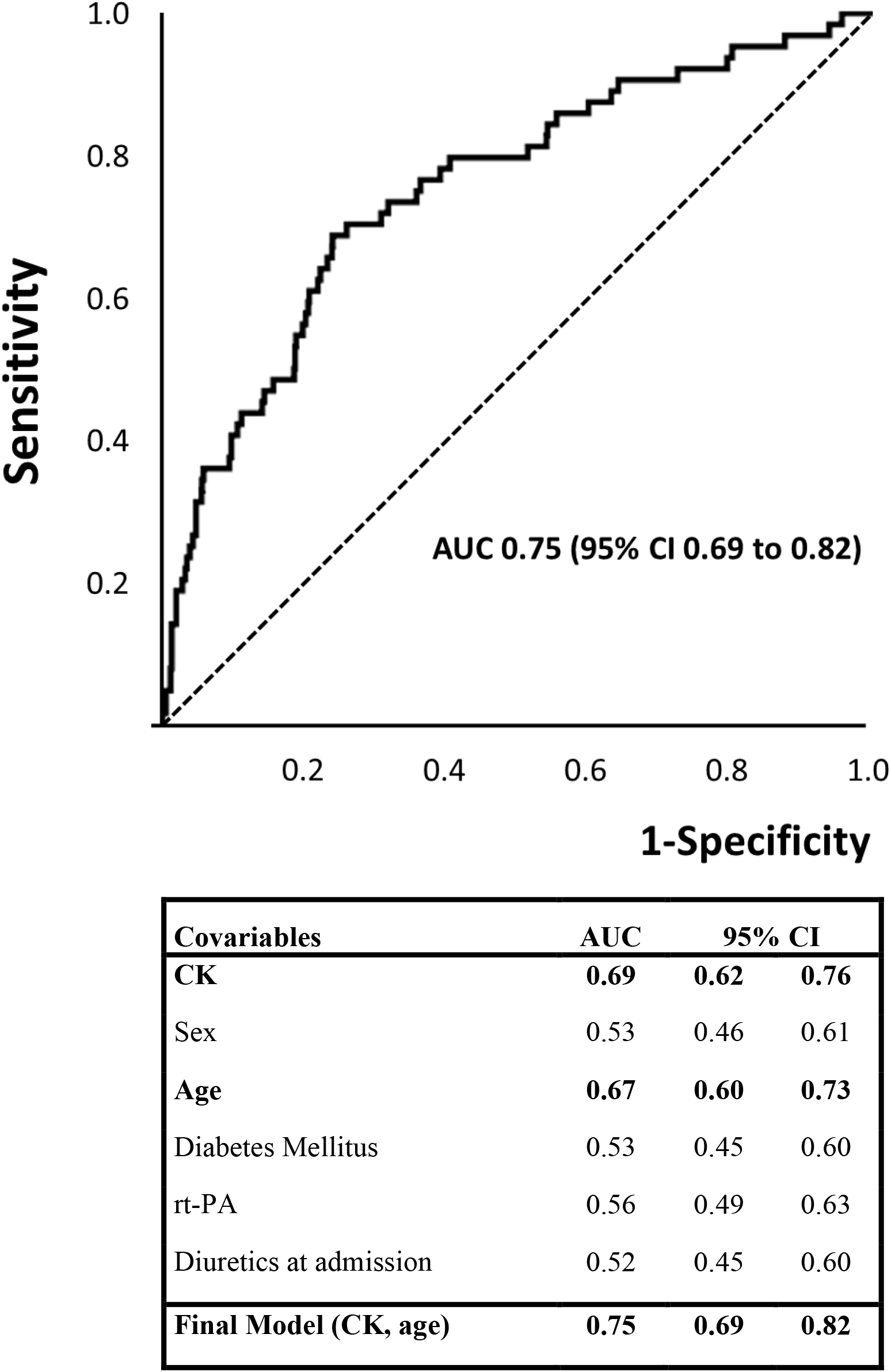
C-indices of covariables and final logistic regression models. AUC, area under the curve; CK, creatine kinase; rt-PA, recombinant tissue-type plasminogen activator use; parameter units are as in Table 1. Panel A. C-index CK and Major Bleeding Panel B. C-index CK and Major Bleeding, Stroke or all-cause mortality

## Discussion

To our knowledge, this is the first report on the association of the ADP-scavenger enzyme CK with bleeding during NSTE-ASC. The data indicate that, consonant with earlier findings in STEMI, (log) CK is independently associated with non-fatal and fatal major bleeding, with a 3-time greater odds compared to the absence of major bleeding and survival in patients treated with thrombolytic or antithrombotic drugs for NSTE-ACS. The association was independent of heparin dose or rt-PA use. Age was another strong, independent risk factor for major bleeding as previously noted.^3^

The biologically plausibility regarding ADP binding (Figure 1), the observed strong, consistent, independent (nonspurious) association with bleeding in large multicenter clinical studies, temporal precedence, the dose-effect relationship, and the experimental evidence on inhibition of platelet aggregation,^6-8^ do not refute the hypothesis that the ADP-scavenging enzyme CK increases bleeding risk. Plasminogen activators, assigned to half of the presented patients, are still in use,^15,16^ and although thrombolysis has been largely replaced by primary percutaneous coronary intervention, (adjunctive) antithrombotic therapy remains strongly associated with major bleeding and subsequent death.^1-4^ It is highly speculative to suggest that even at moderately increased levels, CK acts in synergy with thrombolytic and antithrombotic drugs to facilitate major bleeding in ACS patients with or without myocardial infarction. These data were collected in a randomized clinical trial with high-risk patients excluded, and the main limitation of the current study is the post-hoc analysis. Currently used risk scores for major bleeding, reporting a C-index of around 0.7, do not yet include CK.^1-4^ New clinical analyses are needed to establish whether models including CK more accurately or more simply stratify unselected ACS patients by risk for major bleeding, at low costs.

With the intensification in potency, number of drugs, and duration of antithrombotic therapy for ACS, identifying patients at risk for hemorrhagic complications has become a major safety issue.^1-4^ CK, the ADP-binding enzyme that is near to being declared obsolete in diagnostic cardiology,^1,2,17^ might need to be dusted off and reassessed for its potential to contribute to hemorrhagic complications, and to aid in the selection of patients with the best benefit/risk ratio during contemporary ACS treatment.

## Data Availability

The Manuscript was prepared using TIMI 3 Research Materials obtained from the NHLBI Biologic Specimen and Data Repository Information Coordinating Center. The content does not necessarily reflect the opinions or views of the TIMI 3 study members or the NHLBI.

https://biolincc.nhlbi.nih.gov/

## References

1. Roffi M, Patrono C, Collet JP, Mueller C, Valgimigli M, Andreotti F, Bax JJ, Borger MA, Brotons C, Chew DP, Gencer B, Hasenfuss G, Kjeldsen K, Lancellotti P, Landmesser U, Mehilli J, Mukherjee D, Storey RF, Windecker S; ESC Scientific Document Group. 2015 ESC Guidelines for the management of acute coronary syndromes in patients presenting without persistent ST-segment elevation – 2015 ESC Guidelines for the management of acute coronary syndromes in patients presenting without persistent ST-segment elevation: Task Force for the Management of Acute Coronary Syndromes in Patients Presenting without Persistent ST-Segment Elevation of the European Society of Cardiology (ESC). Eur Heart J. 2016;37:267–315.

2. Levine GN, Bates ER, Bittl JA, Brindis RG, Fihn SD, Fleisher LA, Granger CB, Lange RA, Mack MJ, Mauri L, Mehran R, Mukherjee D, Newby LK, O’Gara PT, Sabatine MS, Smith PK, Smith SC Jr. 2016 ACC/AHA Guideline Focused Update on Duration of Dual Antiplatelet Therapy in Patients With Coronary Artery Disease: A Report of the American College of Cardiology/American Heart Association Task Force on Clinical Practice Guidelines: An Update of the 2011 ACCF/AHA/SCAI Guideline for Percutaneous Coronary Intervention, 2011 ACCF/AHA Guideline for Coronary Artery Bypass Graft Surgery, 2012 ACC/AHA/ACP/AATS/PCNA/SCAI/STS Guideline for the Diagnosis and Management of Patients With Stable Ischemic Heart Disease, 2013 ACCF/AHA Guideline for the Management of ST-Elevation Myocardial Infarction, 2014 AHA/ACC Guideline for the Management of Patients With Non–ST-Elevation Acute Coronary Syndromes, and 2014 ACC/AHA Guideline on Perioperative Cardiovascular Evaluation and Management of Patients Undergoing Noncardiac Surgery. Circulation. 2016;134:e123–e155.

3. Moscucci M, Fox KA, Cannon CP, Klein W, López-Sendón J, Montalescot G, White K, Goldberg RJ. Predictors of major bleeding in acute coronary syndromes: the Global Registry of Acute Coronary Events (GRACE). Eur Heart J. 2003;24:1815–1823.

4. De Silva K, Myat A, Cotton J, James S, Gershlick A, Stone GW. Bleeding associated with the management of acute coronary syndromes. Heart. 2017;103:546–562.

5. Gremmel T, Frelinger AL, Michelson AD. Platelet Physiology. Semin Thromb Hemost. 2016;42:191–204.

6. Brewster LM, Fernand J. Creatine kinase is associated with bleeding after myocardial infarction. MedRxiv. 2019;19012039.

7. Brewster LM. Extracellular creatine kinase may modulate purinergic signalling. Figshare. 2019; 11364932

8. Brewster LM. Creatine kinase, energy reserve, and hypertension: from bench to bedside. Ann Transl Med. 2018;6:292.

9. TIMI IIIA Investigators. Early effects of tissue-type plasminogen activator added to conventional therapy on the culprit coronary lesion in patients presenting with ischemic cardiac pain at rest. Results of the Thrombolysis in Myocardial Ischemia (TIMI IIIA) Trial. Circulation. 1993;87:38–52.

10. TIMI IIIB Investigators. Effects of Tissue Plasminogen Activator and a Comparison of Early Invasive and Conservative Strategies in Unstable Angina and Non-Q-Wave Myocardial Infarction. Results of the TIMI IIIB Trial. Circulation 1994;89:1545–1556.

11. Bovill EG, Terrin ML, Stump DC, Berke AD, Frederick M, Collen D, Feit F, Gore JM, Hillis LD, Lambrew CT, Leiboff R, Mann KG, Markis JE, Pratt CM, Sharkey SW, Sopko G, Tracy RP, Chesebro JH. Hemorrhagic events during therapy with recombinant tissue-type plasminogen activator, heparin, and aspirin for acute myocardial infarction. Results of the Thrombolysis in Myocardial Infarction (TIMI), Phase II Trial. Ann Intern Med. 1991;115:256–65.

12. Hosmer DW, Lemeshow S. Applied logistic regression. 2nd Ed. John Wiley and Sons Inc. New York, 2000, Ch. 8.5, sample size issues when fitting logistic regression models, pp 346–347.

13. Cook NR. Use and misuse of the receiver operating characteristic curve in risk prediction. Circulation 2007;115:928–935.

14. Wasserstein RL, Schirm AL, Lazar NA. Moving to a world beyond “p < 0.05”. Am Stat 2019:73 (sup1):1–19.

15. McCaul M, Lourens A, Kredo T. Pre-hospital versus in-hospital thrombolysis for ST-elevation myocardial infarction. Cochrane Database Syst Rev. 2014;9:CD010191.

16. Powers WJ, Rabinstein AA, Ackerson T, Adeoye OM, Bambakidis NC, Becker K, Biller J, Brown M, Demaerschalk BM, Hoh B, Jauch EC, Kidwell CS, Leslie-Mazwi TM, Ovbiagele B, Scott PA, Sheth KN, Southerland AM, Summers DV, Tirschwell DL; American Heart Association Stroke Council. 2018 Guidelines for the Early Management of Patients With Acute Ischemic Stroke: A Guideline for Healthcare Professionals From the American Heart Association/American Stroke Association. Stroke 2018;49:e46–e110.

17. Alvin MD, Jaffe AS, Ziegelstein RC, Trost JC. Eliminating Creatine Kinase-Myocardial Band Testing in Suspected Acute Coronary Syndrome: A Value-Based Quality Improvement. JAMA Intern Med. 2017;177:1508–1512.

